# Equity and feasibility of remote photoplethysmography for hypertension screening in darker-skinned populations in Nigeria: a multi-site field study

**DOI:** 10.1101/2025.10.13.25337878

**Authors:** David Dasa, Philip Davies

**Affiliations:** School of Computing and Engineering, Bournemouth University, UK

**Keywords:** Remote photoplethysmography (rPPG), Hypertension, Darker skin tones, Facial tribal markings, Usability

## Abstract

**Objective:** To evaluate the feasibility, diagnostic accuracy, and acceptability of a remote photoplethysmography (rPPG) blood pressure screening tool among adults with Fitzpatrick skin types V and VI in Kebbi State, Nigeria, with additional assessment of the influence of facial tribal markings and internet connectivity.

**Methods:** We enrolled 306 patients and 30 staff across three hospitals. rPPG blood pressure estimates (Lifelight app) were compared with OMRON M2 cuff measurements (reference standard; 140/90 mmHg thresholds). Outcomes included the proportion of participants for whom the tool produced a blood pressure estimate, agreement (Bland-Altman), diagnostic accuracy (sensitivity/specificity), and patient/staff perceptions.

**Results:** The tool produced blood pressure estimates for 249 of 306 participants (81.4%), forming the analyzable index-test set. Agreement with the reference standard was limited (SBP MAE 15.37 mmHg, RMSE 19.93; DBP MAE 10.91 mmHg, RMSE 13.61). Within this set, sensitivity for hypertension detection was very low (SBP 0.04; DBP 0.10), with complete failure to detect elevated systolic blood pressure in Fitzpatrick type VI participants (sensitivity = 0.00). Specificity was higher (SBP 0.99; DBP 0.89). Performance was poorest in Fitzpatrick type VI. Lower bandwidth correlated with higher tool failure rates (maximum *r* = −0.69, minimum *r* = −0.51). Tribal markings were not associated with obtaining a reading, but subgroup accuracy differed (e.g., SBP bias −9.97 mmHg with marks). Despite these limitations, 70% of patients rated accuracy favourably and more than 90% of staff expressed willingness to adopt the tool, indicating a perception-performance gap.

**Conclusion:** Remote photoplethysmography was acceptable to patients and staff but is not recommended for population screening without further algorithmic and operational improvements. Despite high specificity (systolic 0.99), critically low sensitivity (systolic 0.04 overall, 0.00 for Fitzpatrick VI) means most hypertensive cases would be missed in screening programmes. Algorithmic refinement for darker skin tones and robust offline or low-bandwidth capability are prerequisites for equitable deployment in low-resource settings.

## 1. Introduction

Hypertension affects ∼31% of adults globally and is a leading cause of cardiovascular morbidity and mortality; nearly half of hypertensive adults are unaware of their condition [1, 2, 3, 4]. The UK National Institute for Health and Care Excellence (NICE) defines clinic hypertension as ≥140/90 mmHg, confirmed by ambulatory/home averages ≥135/85 mmHg [5]. Scaling detection and treatment could avert an estimated 76 million deaths by 2050 [6]. In Nigeria, prevalence is ∼30.6% with substantial hospital burden; prevalence across Africa remains among the highest globally [7, 8, 9]. Conventional cuff measurement depends on trained staff, calibrated devices and time—resources that can be scarce in low-resource settings amid workforce shortages [10, 11, 12, 13].

Remote photoplethysmography (rPPG) infers blood-volume pulse from subtle skin reflectance changes captured by commodity cameras and has been proposed for contactless vital signs estimation [14, 15], the rPPG principle is illustrated in Figure 1. However, training/validation datasets frequently under-represent darker skin, risking performance disparities [16, 17, 18]. For example, MMSE-HR, AFRL and UBFC-rPPG contain ∼10%, 0% and ∼5% dark-skinned subjects, respectively [19, 20, 21, 16]. Large evaluations such as VISION-D/V included very few Fitzpatrick V/VI participants, limiting subgroup inference [22]. Connectivity and deployment constraints in rural clinics further complicate use [23, 24, 25, 26]. Smartphone ubiquity strengthens the case for contactless tools if equity and infrastructure issues are addressed [27].

**Figure 1:**
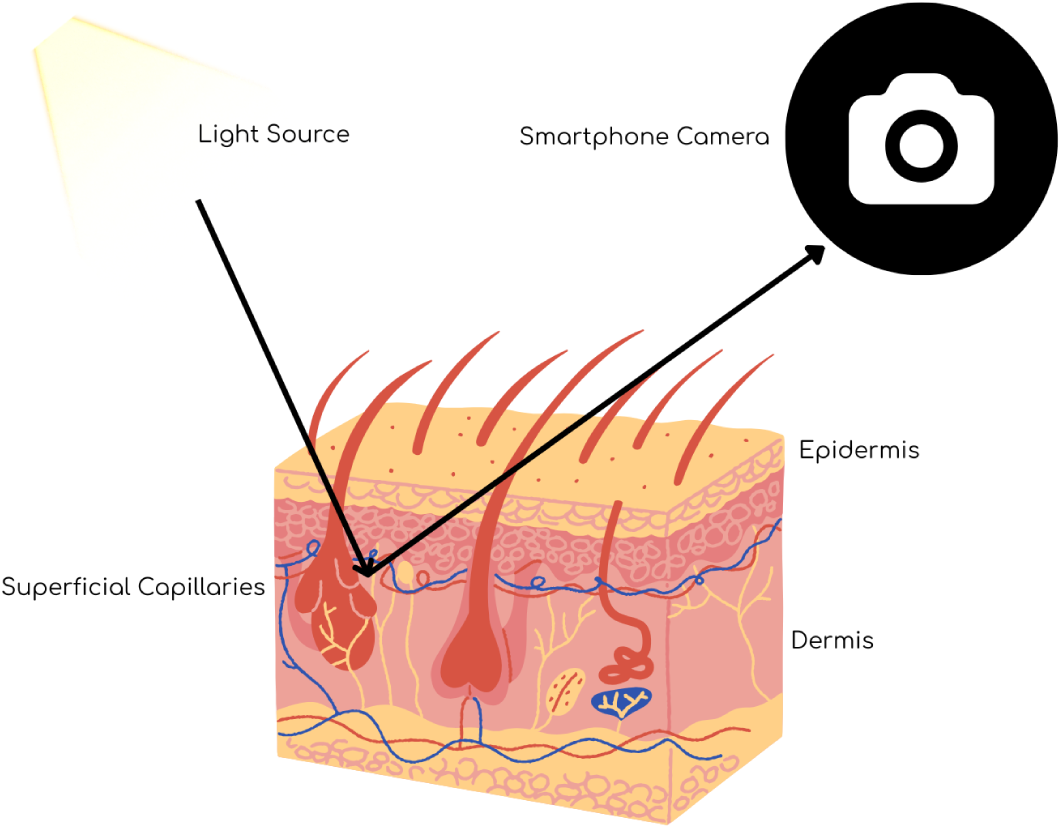
Schematic of rPPG: light–tissue interaction produces small reflected-light variations captured by a camera and processed to estimate pulse/BP. Figure created by the authors, concept adapted from [14].

We conducted a prospective field evaluation of Lifelight (Xim Ltd), an rPPG app that estimates systolic/diastolic blood pressure (SBP/DBP) from a 40 s facial reflection capture, in adults with Fitzpatrick types V–VI in Kebbi State, Nigeria. We assessed feasibility, agreement with a validated cuff reference, diagnostic accuracy at 140/90 mmHg thresholds, and user acceptability, and examined two equity-relevant factors: facial tribal markings and internet bandwidth [28, 29].

### 1.1. rPPG accuracy and limitations in diverse populations

Dataset imbalance and optical differences across skin tones can degrade rPPG performance for under-represented groups [16, 17, 18]. Reported dark-skin proportions in common datasets are low [19, 20, 21, 16]; large Lifelight evaluations reported overall accuracy but had very few V/VI participants, restricting equity conclusions [22]. Comparative signal-extraction work (e.g., ICA/POS/CHROM/PCA) shows no universally superior method, and ML models for BP estimation require diverse data and raise interpretability concerns [30, 31, 32, 33, 34, 35]. Reviews also note small, controlled cohorts and limited real-world testing [36].

### 1.2. Clinical validation of rPPG tools

Emergency-department evaluations have shown promising correlations for some rPPG implementations [37], while VISION-D/V validated Lifelight against standard measurements with limited dark-skin representation [22]. These gaps motivate independent, spectrum-balanced field testing in settings where lighting and connectivity vary [22, 23, 24, 25].

### 1.3. Impact of facial markings on rPPG accuracy

Facial scarification/tattoos (“tribal markings”)—culturally significant in parts of Nigeria—could alter local reflectance and potentially affect rPPG signals; this has not been systematically studied [38, 39, 14]. We therefore recorded the presence/absence of facial tribal markings to explore any measurement effects. Examples of common facial tribal mark patterns are shown in Figure 3.

### 1.4. Identified gaps and study objectives

We addressed four gaps: (i) under-representation of darker skin in rPPG datasets, (ii) limited independent field validation, (iii) unknown impact of facial markings, and (iv) acceptability in low-resource settings [16, 22, 36, 25, 24]. Accordingly, we evaluated Lifelight’s performance/acceptability in Fitzpatrick V/VI adults in Nigeria, including effects of markings and rural deployment barriers.

## 2. Methods

### 2.1. Design, setting and participants

We performed a prospective observational field study across three hospitals (two urban, one rural) in Kebbi State, Nigeria (13 June–2 July 2024), Kebbi State’s location is shown in Figure 2. Adults (≥18 y) with Fitzpatrick V or VI were eligible; exclusions were inability to consent, pregnancy, relevant facial dermatologic conditions, heavy makeup, and inability to remain still for ∼40 s. Two cohorts were enrolled: patients (primary feasibility/accuracy) and staff (acceptability). Convenience sampling was used.

**Figure 2:**
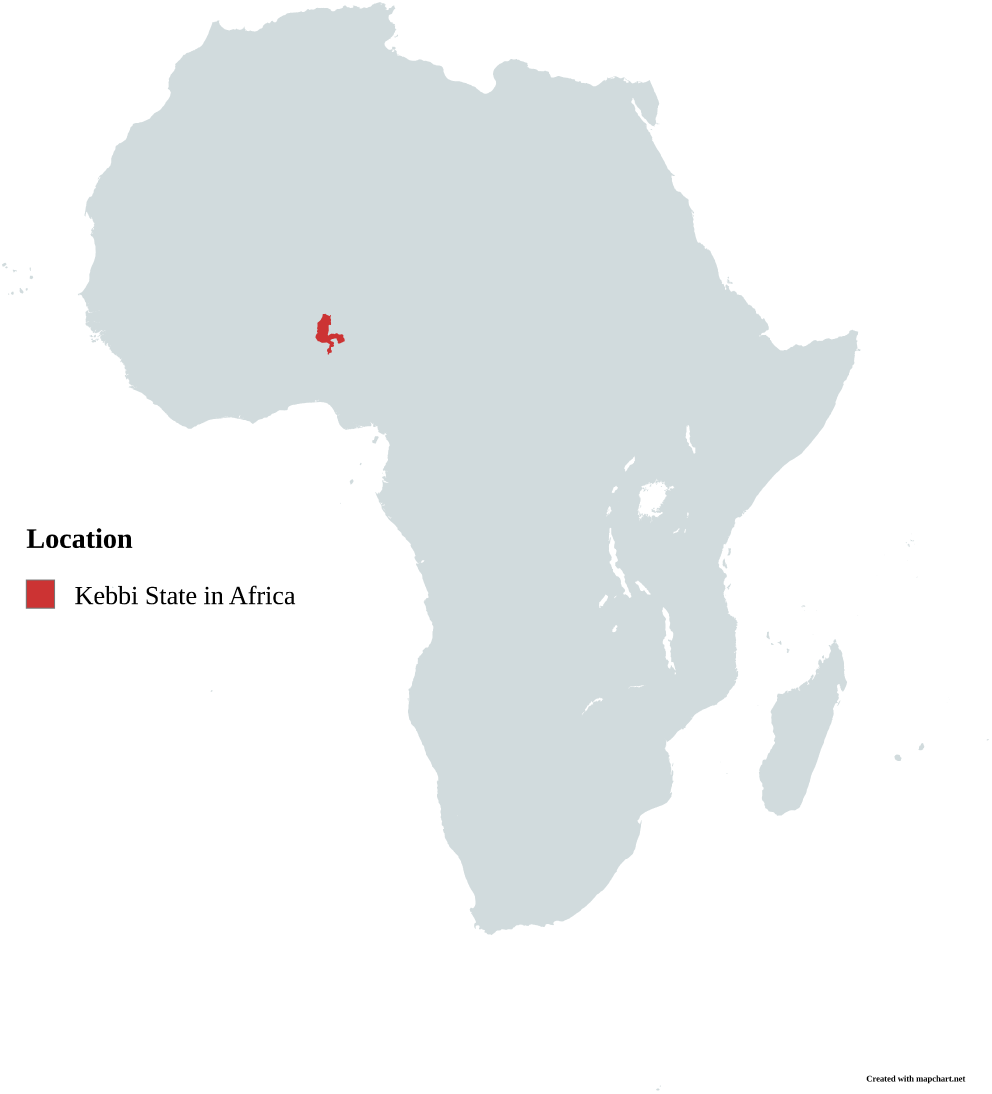
Map of Nigeria with Kebbi State highlighted. Map created by the authors using mapchart.net.

### 2.2. Devices and procedure

OMRON M2 automated cuffs (Boots^TM^ branded) procedures followed ANSI/AAMI/ISO 81060-2 clinical investigation guidance [28, 40]. Lifelight (LLTry v2.0.0; Xim Ltd) ran on an iPad (8th gen; iPadOS 17.5.1; front camera 1.2 MP) [41]. Participants rested 5 min seated before measurements [42]. Sequence: two OMRON readings (2 min apart), a 40 s Lifelight capture (up to three attempts), then a final OMRON reading to align temporally with the index test. Distance/lighting were standardised where possible; a ring light was used if ambient light was insufficient. Lifelight is CE-marked software with MHRA registration and requires stable connectivity on supported devices [43, 26, 23, 44, 45].

### 2.3. Ascertainment of skin type and facial tribal markings

Skin type was assessed in-person by trained healthcare staff using a printed Fitzpatrick visual chart [46, 47]. Facial tribal markings were documented as a binary variable (present/absent) following non-invasive visual inspection during enrolment. Examples of common facial tribal mark patterns are shown in Figure 3. No photographs or videos of participants were captured by the application, and only non-identifying biodata summaries were stored in accordance with national and international data protection guidelines [48, 49].

**Figure 3:**
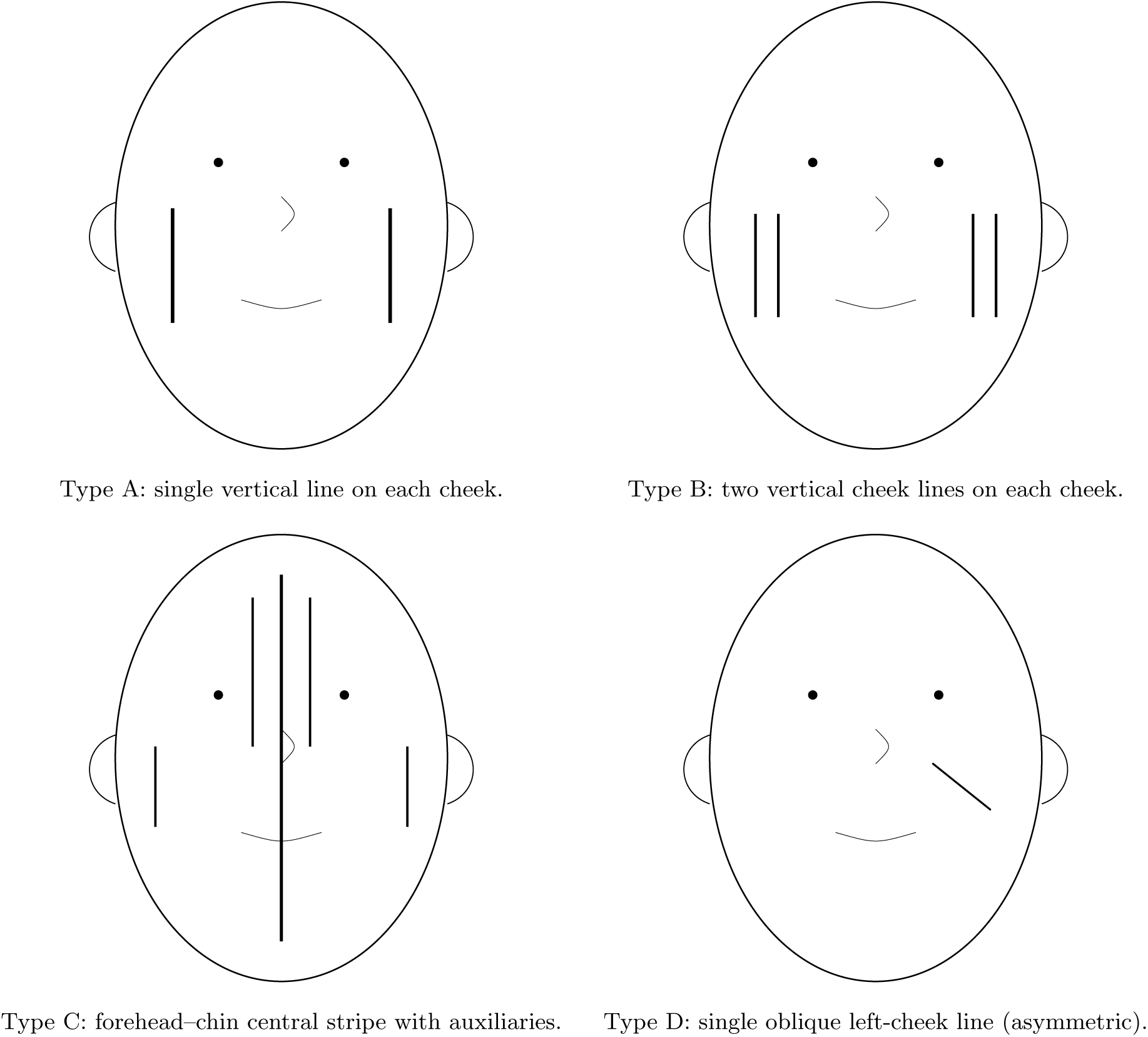
Schematic, non-identifying examples of facial tribal markings (A–D illustrate patterns). Many international readers may be unfamiliar with culturally specific facial scarification/tattooing; these neutral line-art illustrations are provided as an explanatory, nonidentifying alternative to participant photographs. In the study markings were recorded only as a binary variable (present / absent) and were not sub-typed. [38, 39]

### 2.4. Reference standard and analysis sets (STARD)

This diagnostic-accuracy study follows STARD guidance [29]. For hypertension classification, the reference standard was the mean of three OMRON readings (SBP≥140 and/or DBP≥90 mmHg). For paired continuous comparisons, the final OMRON reading was time-aligned to the Lifelight capture. Participants without a valid Lifelight estimate after ≤3 attempts contributed to feasibility outcomes but were excluded from diagnostic-accuracy analyses.

### 2.5. Outcomes

Primary feasibility outcome: proportion producing an app BP estimate. Agreement outcomes: MAE, RMSE, and Bland–Altman bias and 95% limits of agreement (LoA) for SBP/DBP [50]. Diagnostic outcomes: sensitivity, specificity, PPV and NPV at 140/90 mmHg. Secondary outcomes: subgroup performance (Fitzpatrick V vs VI; tribal markings present vs absent), bandwidth effects, and acceptability from patient/staff questionnaires.

### 2.6. Connectivity monitoring and site characteristics

Bandwidth (MTN Nigeria) was measured twice daily (12:00, 16:00) at each site using fast.com; daily minima/maxima were linked to session-level success/failure and error metrics. Urban/rural contexts are shown in Figure 2 [25, 24].

### 2.7. Participant workflow, questionnaires, and staff training

Eligibility was confirmed, written informed consent obtained (English/Hausa; back-translated), and demographics recorded. Mid–upper-arm circumference was used solely for cuff selection; MUAC/BMI were not recorded. Participants sat ∼50 cm from the iPad on a tripod facing natural light; the app prompted stillness. Staff received a one-day training on consent, skin-type assignment, tribal-mark ascertainment, positioning/lighting and troubleshooting. Fitzpatrick skin typing was performed using standardized visual assessment against reference photographic charts. Patient and staff questionnaires (developed from study objectives and digital-health usability literature; clinician-reviewed; translated/back-translated) captured perceived accuracy, comfort, workflow impact, barriers and willingness to adopt [51].

### 2.8. Data management, privacy and security

Paper CRFs were stored in locked cabinets; data were transcribed to CSV and stored in a password-protected OneDrive accessible only to study staff. Identifiers were replaced with study IDs prior to analysis. In line with protocol and regulation, individual-level raw videos were deleted after preprocessing; only de-identified summaries/aggregates were retained [48, 52, 53, 49].

### 2.9. Statistical analysis

Continuous agreement was summarised with paired differences, MAE/RMSE, and Bland–Altman plots with nonparametric bootstrap 95% CIs for bias and LoA [50]. Pearson correlations with the reference were reported where relevant. Diagnostic accuracy used exact (Clopper–Pearson) 95% CIs for sensitivity/specificity/PPV/NPV and Wilson CIs for accuracy [54, 55]. Subgroup estimates were interpreted cautiously given small numerators. Analyses were performed in Python (`pandas` [56], `NumPy` [57], `SciPy` [58], `statsmodels` [59]); plots were generated using `matplotlib` [60] and `seaborn` [61].

### 2.10. Sample size and power

Target enrolment (patients *n* = 306; staff *n* = 30) exceeded simple requirements for paired mean comparisons and permitted prespecified subgroup summaries. A paired *t*-test calculation (two-sided *α* = 0.05, 80% power) indicates *n* ≈ 34 to detect a medium effect (Cohen’s *d* = 0.5); regression guidance of 10–20 observations per predictor suggests *n* ≥ 120 for modest multivariable models [62, 63]. Precision for subgroup sensitivities was expected to be limited; exact CIs are therefore reported.

### 2.11. STARD flow and handling of indeterminate results

Participants with repeated Lifelight failures (up to three attempts) were classified as *failed/dropped* for the index test and excluded from diagnostic-accuracy analyses; they contributed to feasibility outcomes. Participant flow is shown in Figure 4 [29].

**Figure 4:**
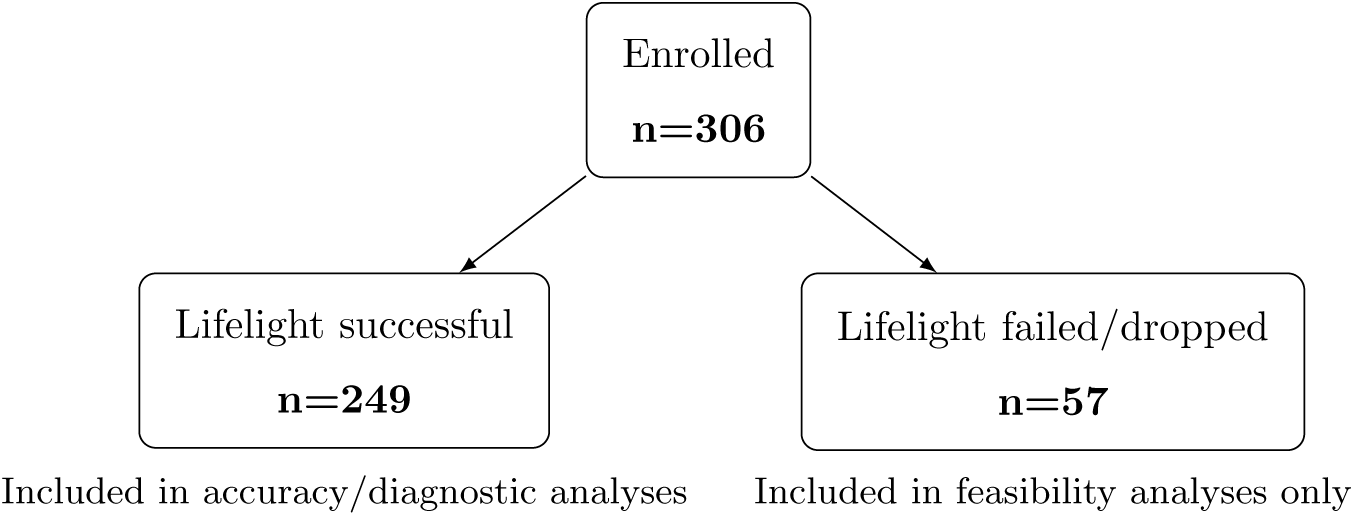
Participant flow for index (Lifelight) and reference (OMRON) testing (STARD).

### 2.12. Ethics

Ethics approval: Kebbi State Ministry of Health (107:017/2024) and Bournemouth University (55993). Written informed consent was obtained from each participant. Incidental hypertension was referred per local care pathways.

## 3. Results

### 3.1. Cohort, demographics and feasibility

We enrolled 306 patients and 30 staff. Of 306 patients, 287 completed questionnaires (response rate 93.8%); not all respondents answered every item. Among those declaring sex, 149 were female and 137 male. Age distribution is shown in Table 1; most were 18–30 y (n=133), with 78 aged 31–45 y, 45 aged 46–60 y, and 30 aged *>*60 y. By clinical assessment, Fitzpatrick type V was more prevalent (174/306; 56.9%) than type VI (132/306; 43.1%). Facial tribal markings were present in 88/306 (28.7%).

**Table 1:** Age distribution of participants with available data.

| Age group (years) | Number of participants |
| --- | --- |
| 18–30 | 133 |
| 31–45 | 78 |
| 46–60 | 45 |
| >60 | 30 |
| Total | 286 |

Lifelight *produced a BP estimate* for 249/306 patients (81.4%); 57 failed/dropped (see STARD flow). Urban sites achieved higher success than the rural site, where no valid readings were obtained (Figure 5). Within the successful-read analysis set (*n* = 249), OMRON identified 70/249 (28.1%) hypertensives (Table 3).

**Figure 5:**
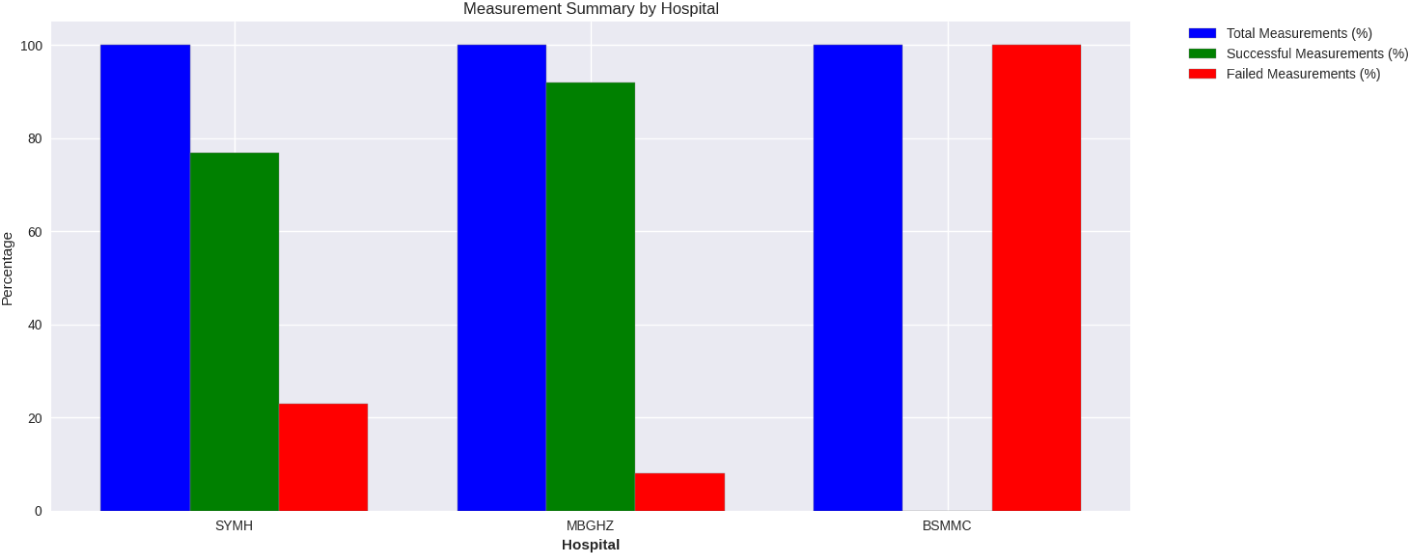
Successful Lifelight measurements by study centre: two urban sites vs one rural site.

### 3.2. Connectivity and feasibility

Daily bandwidth correlated negatively with failure rates (max speed *r* = −0.69; min speed *r* = −0.51; Figure 6), consistent with the rural site’s 100% failure (Table 2).

**Figure 6:**
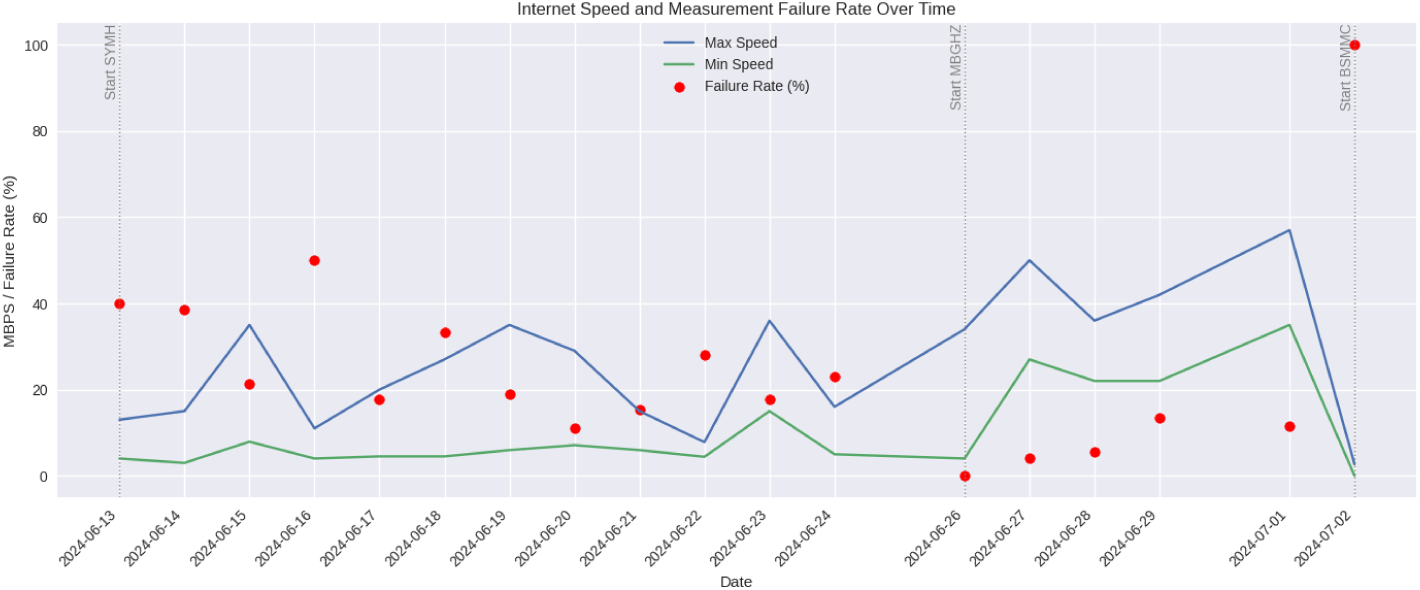
Daily internet speeds (min/max) vs Lifelight failure rates.

**Table 2:** Session duration, counts, throughput and failure rates by site.

| Site | Total hours | Total measurements | Failed measurements | Measurements/hour | Failure rate (%) |
| --- | --- | --- | --- | --- | --- |
| SYMH | 80 | 203 | 47 | 2.54 | 23.20 |
| MBGHZ | 36 | 100 | 8 | 2.78 | 8.0 |
| BSMMC | 4 | 2 | 2 | 0.50 | 100.0 |

Correlations between bandwidth and absolute error were weak; connectivity primarily affected the *probability of obtaining a reading* rather than error magnitude.

### 3.3. Factors associated with measurement success

We first examined whether the physiological level of blood pressure itself predicted whether Lifelight would return a reading. Visual inspection of scatter and distribution plots showed substantial overlap between retained and dropped attempts, with no clustering of failures at extreme SBP/DBP values (Figures 7 and 8). These impressions were borne out by contingency analyses (Table 3), which found no association between elevated BP (by the reference) and success (all *p >* 0.34; negligible Cramer’s *V* ).

**Figure 7:**
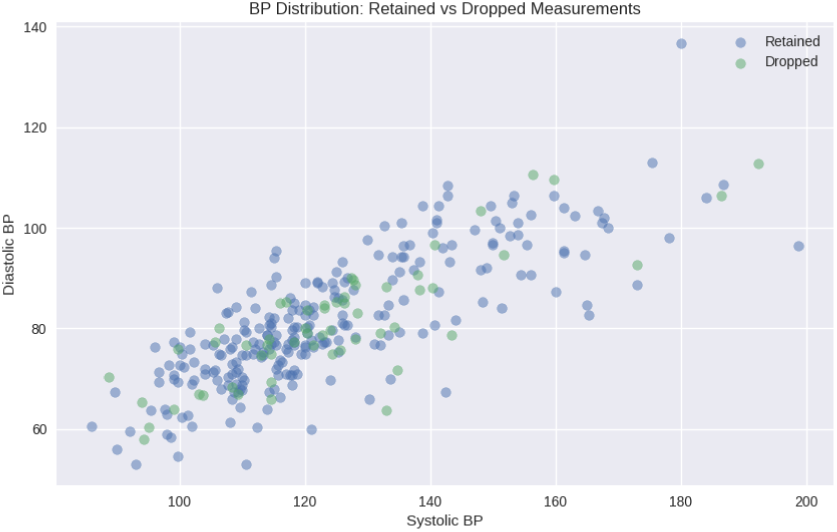
Scatter of SBP/DBP for retained vs dropped attempts. No clustering at extremes among failures.

**Figure 8:**
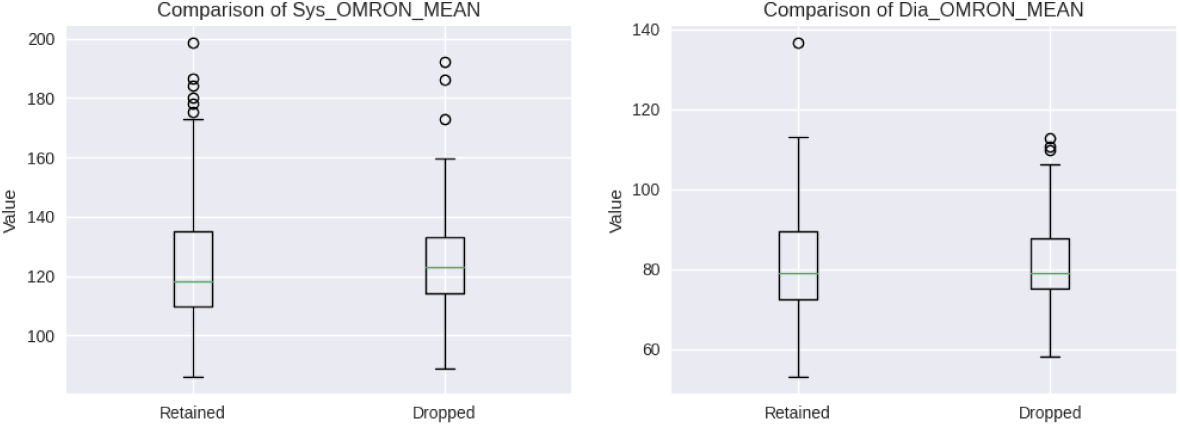
Boxplots of SBP/DBP by retained vs dropped attempts, showing overlapping distributions.

**Table 3:** Success vs failure by BP status (reference standard).

| Status | Systolic BP |  | Diastolic BP |  |
| --- | --- | --- | --- | --- |
|  | Successful | Failed | Successful | Failed |
| Non-elevated | 197 | 47 | 188 | 47 |
| Elevated | 52 | 10 | 61 | 10 |

To complement these point clouds, we compared central tendencies and spread. The boxplots likewise suggest that measurement success was not driven by baseline BP level (Figure 8).

We then turned to participant characteristics. Table 3 summarises success/failure by BP category for reference; in contrast, skin type showed a clear association with success (Table 4; *χ*^2^ = 22.25, *p* = 2.39 × 10^−6^; Cramer’s *V* = 0.27), with more failures in type VI. The presence of facial tribal markings did *not* affect success (Table 5; *χ*^2^ = 0.0012, *p* = 0.9721).

**Table 4:**
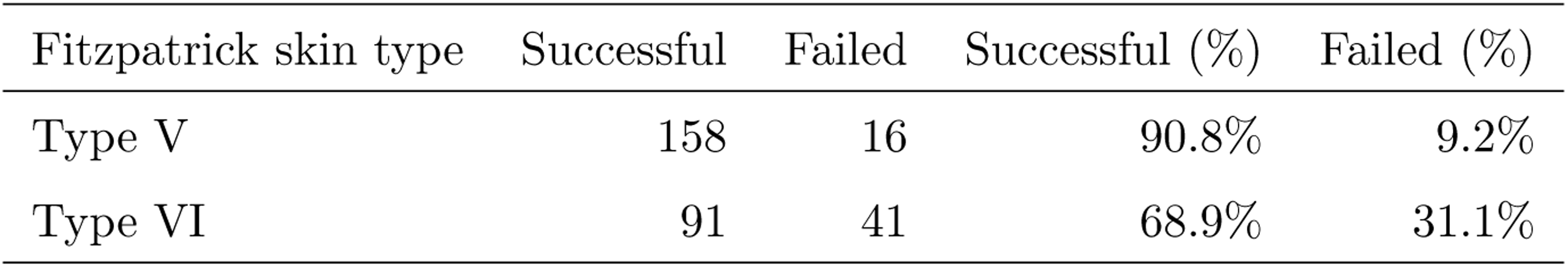
Measurement success by Fitzpatrick type. Percentages are row-wise (success/failed of that Fitzpatrick group).

**Table 5:** Measurement success by tribal markings (present/absent). Percentages are row-wise.

| Facial tribal markings | Successful | Failed | Successful (%) | Failed (%) |
| --- | --- | --- | --- | --- |
| Present | 71 | 17 | 80.7% | 19.3% |
| Absent | 178 | 40 | 81.7% | 18.3% |

The skin-type contingency table (Table 4) highlights the imbalance in failures between types V and VI.

Finally, success rates were similar irrespective of whether facial tribal markings were present (Table 5); success was 80.7% with markings versus 81.7% without.

### 3.4. Agreement (continuous BP)

Having established which factors were linked to success, we evaluated continuous agreement between Lifelight and the reference. Overall error was substantial (SBP MAE 15.37 mmHg; DBP MAE 10.91 mmHg), and Bland–Altman analyses showed near-zero mean SBP bias but very wide limits of agreement (LoA ∼ ±40 mmHg), with a small positive bias and wide LoA for DBP (Tables 13, 14). Figures 9 and 10 display subgroup patterns: agreement appeared poorest for type VI, and participants with tribal markings showed greater SBP underestimation than those without.

**Figure 9:**
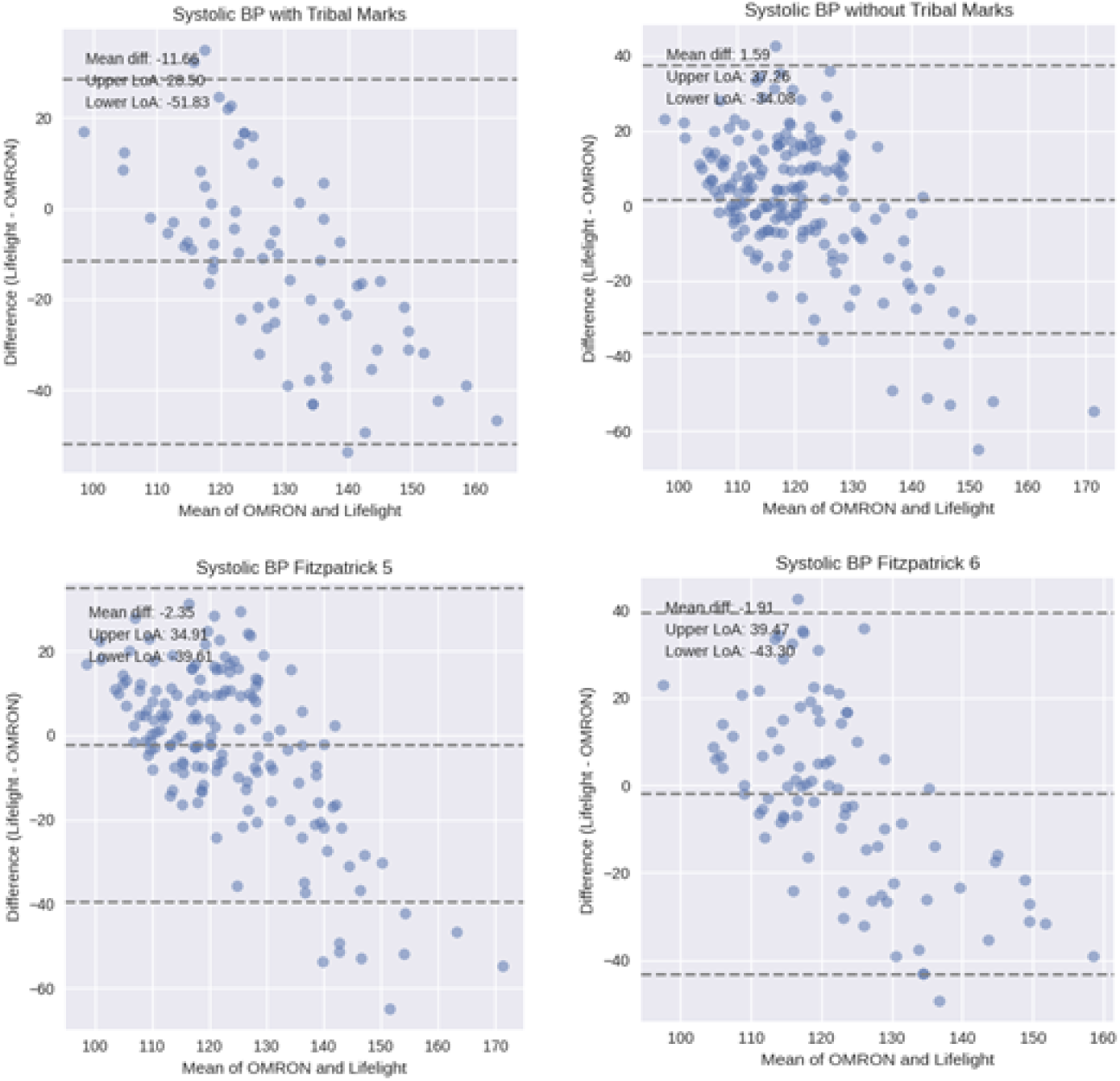
Bland–Altman for **SBP** by subgroup. Agreement poorest for Fitzpatrick VI. Tribal markings were not associated with overall success/failure.

**Figure 10:**
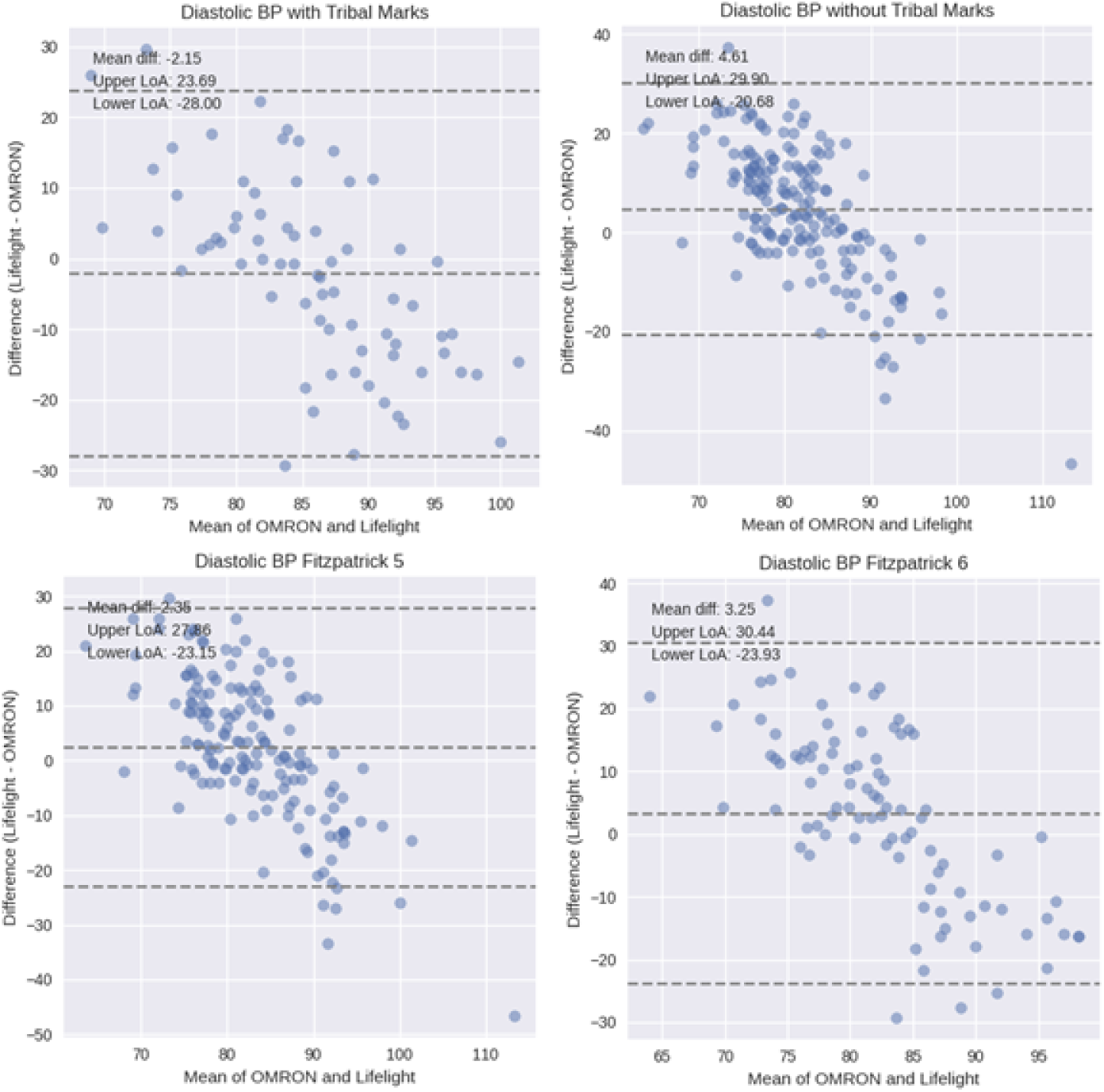
Bland–Altman for **DBP** by subgroup. Positive bias and wide LoA overall.

The diastolic plots similarly indicate wide LoA and a small positive bias overall (Figure 10).

### 3.5. Diagnostic accuracy

We next assessed threshold-based classification. Sensitivity for hypertension detection was very low whereas specificity was higher (Table 6). To make the operating characteristics transparent, we show the overall summary first, then the underlying 2 × 2 tables and, separately, the derived metrics for SBP and DBP.

**Table 6:**
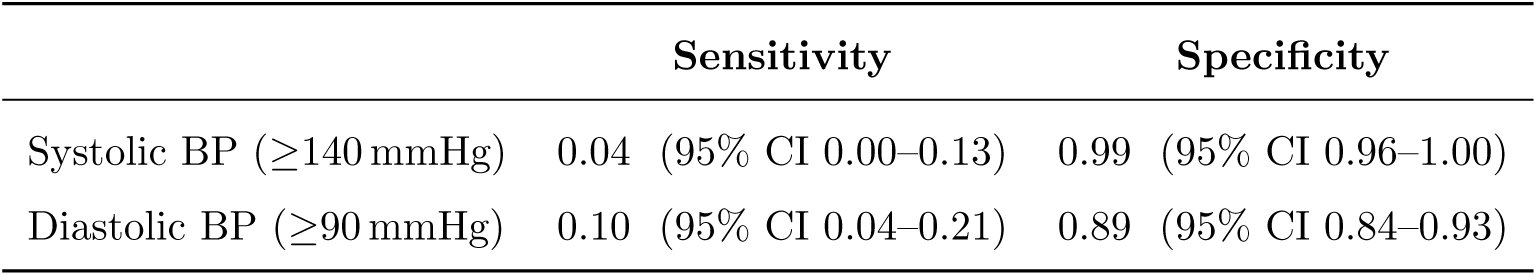
Overall sensitivity and specificity (successful reads only; *n* = 249). Exact (Clopper– Pearson) CIs.

|  | Sensitivity | Specificity |
| --- | --- | --- |
| Systolic BP ($\geq 140$ mmHg) | 0.04 (95% CI 0.00–0.13) | 0.99 (95% CI 0.96–1.00) |
| Diastolic BP ($\geq 90$ mmHg) | 0.10 (95% CI 0.04–0.21) | 0.89 (95% CI 0.84–0.93) |

#### Systolic BP

The confusion matrix below (Table 7) shows that very few true positives drive the low sensitivity.

**Table 7:** Confusion matrix for **SBP** within successful reads (*n* = 249).

| Systolic (SBP $\geq 140$) | OMRON + | OMRON – | Total |
| --- | --- | --- | --- |
| **Lifelight** + | TP = 2 | FP = 2 | 4 |
| **Lifelight** – | FN = 50 | TN = 195 | 245 |
| Total | 52 | 197 | 249 |

Derived operating characteristics for SBP are reported next for completeness.

#### Diastolic BP

DBP shows slightly higher sensitivity but reduced specificity due to more false positives, as the confusion matrix indicates (Table 9).

**Table 8:** Diagnostic performance estimates for **SBP** (successful reads; *n* = 249).

| Metric | Estimate | 95% CI |
| --- | --- | --- |
| Sensitivity | 0.04 | 0.00–0.13 |
| Specificity | 0.99 | 0.96–1.00 |
| PPV | 0.50 | 0.07–0.93 |
| NPV | 0.80 | 0.74–0.84 |
| Accuracy | 0.79 | 0.74–0.84 |

**Table 9:**
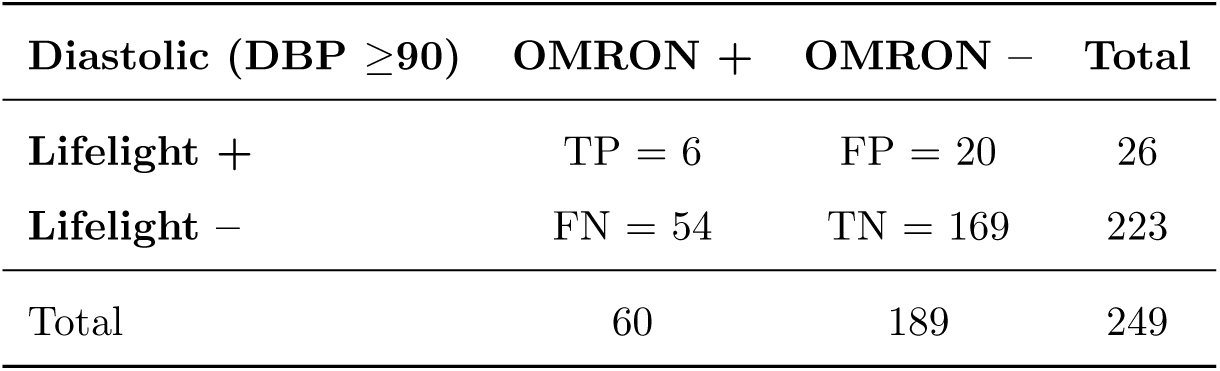
Confusion matrix for **DBP** within successful reads (*n* = 249).

| Diastolic (**DBP** $\geq 90$) | OMRON + | OMRON - | Total |
| --- | --- | --- | --- |
| **Lifelight** + | TP = 6 | FP = 20 | 26 |
| **Lifelight** - | FN = 54 | TN = 169 | 223 |
| Total | 60 | 189 | 249 |

The corresponding DBP metrics are shown below to mirror the SBP presentation.

### 3.6. Subgroups

We then explored whether performance varied by facial phenotype and skin tone. The raw counts underpinning subgroup estimates are shown in Table 11; they illustrate the tendency toward missed hypertensives, especially for SBP and in type VI. The critically low sensitivity for Fitzpatrick type VI (systolic 0.00, 95% CI 0.00-0.16) reflects complete failure to detect any of the 21 hypertensive cases identified by OMRON in this subgroup.

**Table 10:** Diagnostic performance estimates for **DBP** (successful reads; *n* = 249).

| Metric | Estimate | 95% CI |
| --- | --- | --- |
| Sensitivity | 0.10 | 0.04–0.21 |
| Specificity | 0.89 | 0.84–0.93 |
| PPV | 0.23 | 0.09–0.44 |
| NPV | 0.76 | 0.70–0.81 |
| Accuracy | 0.70 | 0.64–0.76 |

**Table 11:** Counts identified with elevated BP by OMRON vs Lifelight across subgroups (successful-read set).

| Subgroup | High BP (any) | Elevated SBP | Elevated DBP | Elevated Both |
| --- | --- | --- | --- | --- |
| OMRON with tribal marks | 36 | 29 | 29 | 22 |
| Lifelight with tribal marks | 11 | 0 | 11 | 0 |
| OMRON without tribal marks | 34 | 23 | 31 | 20 |
| Lifelight without tribal marks | 18 | 4 | 15 | 1 |
| OMRON Fitzpatrick V | 44 | 31 | 36 | 23 |
| Lifelight Fitzpatrick V | 17 | 3 | 15 | 1 |
| OMRON Fitzpatrick VI | 26 | 21 | 24 | 19 |
| Lifelight Fitzpatrick VI | 12 | 1 | 11 | 0 |

Point estimates of sensitivity and specificity (Table 12) confirm consistently low sensitivity and high SBP specificity across strata, with the weakest sensitivities in type VI.

**Table 12:** Point estimates of sensitivity/specificity by subgroup (successful reads only).

| Subgroup / Measure | Sensitivity | Specificity |
| --- | --- | --- |
| **With facial tribal markings** |  |  |
| Systolic BP | 0.04 | 1.00 |
| Diastolic BP | 0.14 | 0.83 |
| **Without facial tribal markings** |  |  |
| Systolic BP | 0.09 | 0.99 |
| Diastolic BP | 0.06 | 0.91 |
| **Fitzpatrick type V** |  |  |
| Systolic BP | 0.06 | 0.99 |
| Diastolic BP | 0.11 | 0.91 |
| **Fitzpatrick type VI** |  |  |
| Systolic BP | 0.00 | 0.99 |
| Diastolic BP | 0.08 | 0.87 |

Exact CIs (Tables 13 and 14) indicate considerable imprecision around sensitivity—particularly in type VI—reinforcing the need for larger, spectrum-balanced samples. We first present SBP results.

**Table 13:** Subgroup diagnostic performance for **SBP** with exact 95% CIs.

| Subgroup (SBP $\geq 140$ mmHg) | Sensitivity (95% CI) | Specificity (95% CI) |
| --- | --- | --- |
| With facial tribal marks (n=71) | 0.04 (0.00–0.18) | 1.00 (0.92–1.00) |
| Without facial tribal marks (n=178) | 0.09 (0.01–0.28) | 0.99 (0.96–1.00) |
| Fitzpatrick type V (n=158) | 0.06 (0.01–0.21) | 0.99 (0.96–1.00) |
| Fitzpatrick type VI (n=91) | 0.00 (0.00–0.16) | 0.99 (0.92–1.00) |

**Table 14:** Subgroup diagnostic performance for **DBP** with exact 95% CIs.

| Subgroup (DBP $\geq 90$ mmHg) | Sensitivity (95% CI) | Specificity (95% CI) |
| --- | --- | --- |
| With facial tribal marks (n=71) | 0.14 (0.04–0.32) | 0.83 (0.69–0.93) |
| Without facial tribal marks (n=178) | 0.06 (0.01–0.21) | 0.91 (0.85–0.95) |
| Fitzpatrick type V (n=158) | 0.11 (0.03–0.26) | 0.91 (0.84–0.95) |
| Fitzpatrick type VI (n=91) | 0.08 (0.01–0.27) | 0.87 (0.76–0.94) |

SBP findings show uniformly high specificity but very low sensitivity across strata; the upper CI bound for type VI remains low (0.16), underscoring risk of missed hypertensives. We next report DBP, where specificity typically falls.

### 3.7. Bandwidth and measurement error

Because bandwidth mainly governed feasibility, we checked whether it also shifted error magnitudes. Correlations with absolute differences were small and inconsistent (Table 15), consistent with a threshold effect: adequate speed prevents failure, but extra bandwidth adds little accuracy.

**Table 15:** Correlations between BP differences and internet speeds.

| BP difference (index-reference) | vs Max speed | vs Min speed |
| --- | --- | --- |
| Systolic | 0.15 | 0.24 |
| Diastolic | 0.10 | 0.09 |

### 3.8. User perceptions

Survey responses provide context for deployment constraints and opportunities. Patients most often flagged connectivity and accuracy as concerns (Figure 11), despite generally favourable comfort and experience ratings.

**Figure 11:**
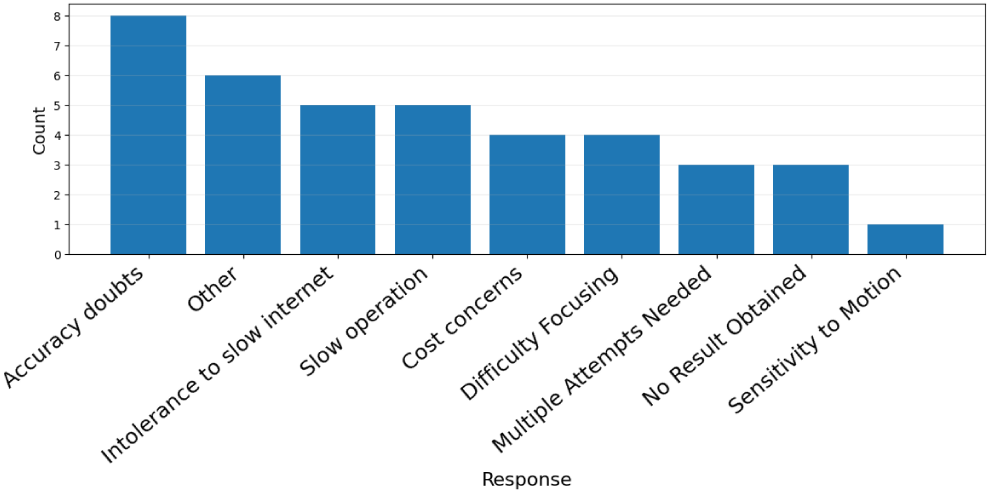
Patient-reported concerns; internet connectivity and accuracy predominated.

The staff cadre mix (Table 16) was predominantly nursing, reflecting the target user group for routine vitals collection.

**Table 16:**
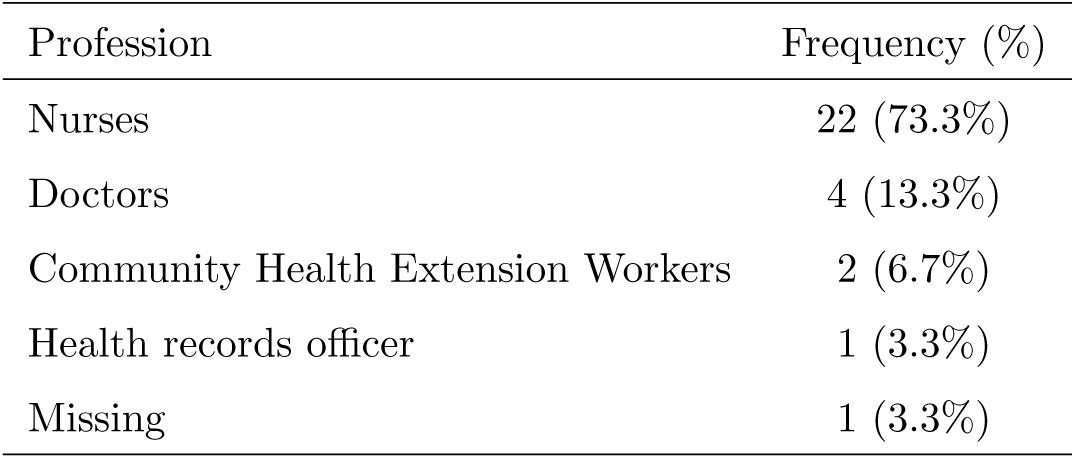
Staff participants by profession (*n* = 30).

| Profession | Frequency (%) |
| --- | --- |
| Nurses | 22 (73.3%) |
| Doctors | 4 (13.3%) |
| Community Health Extension Workers | 2 (6.7%) |
| Health records officer | 1 (3.3%) |
| Missing | 1 (3.3%) |

Perceptions varied by seniority, with more positive views among junior staff (Figure 12); the headline response rates are summarised afterwards.

**Figure 12:**
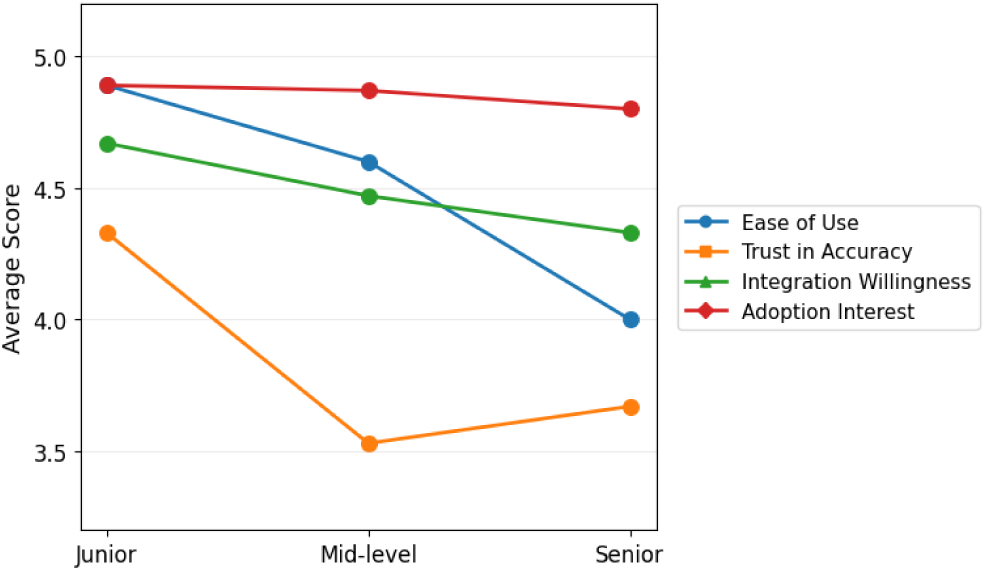
Staff perceptions by years of experience: more positive among junior staff.

Summary metrics (Table 17) align with the quantitative results: high usability and willingness to adopt, tempered by accuracy and connectivity concerns.

**Table 17:**
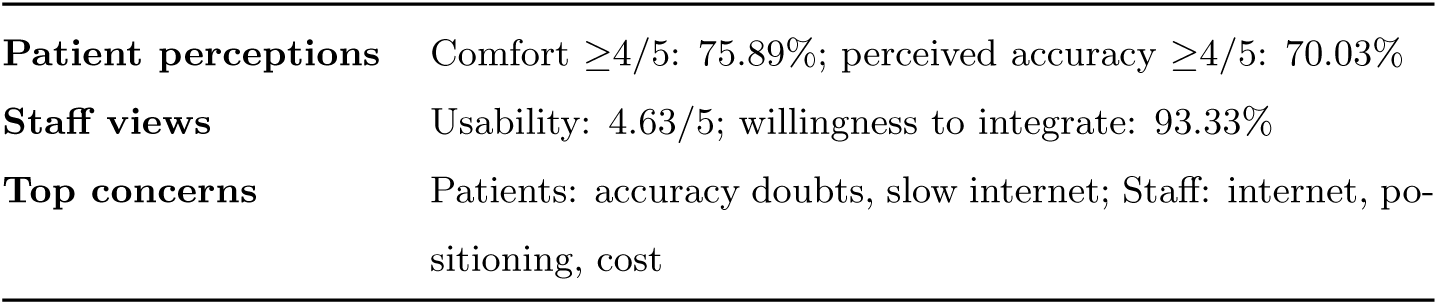
Summary of questionnaire responses.

### 3.9. Summary of key findings

Across sites, Lifelight operated for most urban participants but failed at the rural site due to bandwidth. Key quantitative indicators are summarised in Table 18. Agreement with cuff measurements was weak (SBP MAE 15.37; DBP MAE 10.91; wide LoA), and diagnostic sensitivity was very low, particularly for Fitzpatrick type VI. Specificity was high for SBP but lower for DBP. Connectivity predominantly affected feasibility rather than error magnitude.

**Table 18:** Key quantitative indicators.

|  |  |
| --- | --- |
| **Accuracy** | SBP MAE 15.37, RMSE 19.93; DBP MAE 10.91, RMSE 13.61 |
| **Screening** | Sensitivity: SBP 0.04, DBP 0.10; Specificity: SBP 0.99, DBP 0.89 |
| **Subgroups** | Type VI: SBP sens 0.00, DBP sens 0.08; tribal marks: SBP bias $-9.97$ mmHg |
| **Connectivity** | Failure correlated with low bandwidth (max $r = -0.69$, min $r = -0.51$); weak links to error |
| **Sites** | Urban success 77–92%; rural success 0% |

## 4. Discussion

In a real-world, multicentre evaluation among adults with Fitzpatrick V–VI skin, Lifelight produced valid readings for 81.4% of participants but showed poor agreement with a validated cuff (SBP MAE 15.37 mmHg; DBP MAE 10.91 mmHg; wide LoA) and very low sensitivity for hypertension detection (SBP 0.04; DBP 0.10). High specificity—especially for SBP (0.99)—does not offset the risk of missed hypertensives, which was most pronounced in type VI (SBP sensitivity 0.00). Connectivity primarily affected feasibility (strong negative correlations between bandwidth and failure), with negligible association with absolute error, underscoring the need for offline or edge operation in low-bandwidth settings.

Facial tribal markings did not reduce the probability of obtaining a reading, but subgroup analyses suggested greater SBP underestimation and lower DBP specificity, indicating phenotype-linked optical variability that warrants explicit handling. These findings contrast with earlier evaluations (e.g., VISION-D/V), in which types V/VI were minimally represented [22], and align with concerns that under-representation of darker skin tones can degrade rPPG performance [18].

Strengths include prospective field testing across urban/rural sites, explicit STARD reporting, and integration of quantitative and qualitative data. Limitations include convenience sampling from a single state, a short study window, lack of lux/melanin instrumentation, and no comparator cohort with Fitzpatrick I–IV; subgroup estimates have wide CIs due to small numerators. Overall, the results indicate that current performance is inadequate for screening in similar populations and contexts.

## 5. Conclusion and Future Work

In this independent field evaluation, Lifelight did not meet the sensitivity or agreement thresholds required for hypertension screening. In Fitzpatrick V–VI populations, feasibility was bandwidth-dependent (urban success vs. rural failure) and diagnostic sensitivity was very low—particularly in type VI (SBP sensitivity = 0.00; overall SBP/DBP 0.04*/*0.10)—despite high SBP specificity (0.99). Individual-level agreement with a validated cuff was poor (wide LoA), implying clinically meaningful error even when mean bias is small. Facial tribal markings did not reduce the chance of obtaining a reading, but subgroup bias/specificity patterns suggest additional optical heterogeneity. These findings pertain to this Lifelight research build; other rPPG products require independent, spectrum-balanced validation before generalisation.

### Immediate implications

Until sensitivity and LoA targets are met and independently verified, this app should *not* be used for population hypertension screening in darker-skin–predominant settings. If piloted for any purpose, restrict to a *rule-in* workflow with mandatory cuff confirmation; negative results must not preclude standard BP measurement. Connectivity constraints argue for reliable on-device inference with graceful degradation. These guardrails align with device evaluation and procurement guidance [28, 23, 64].

### Priorities for improvement

- **Algorithmic accuracy:** retrain/calibrate with balanced V/VI representation and diverse facial phenotypes (including scarification); incorporate melanin-aware signal models and augmentation [18].
- **Spectral/engineering robustness:** assess multi-band/IR strategies and skin-tone–adaptive channel weighting to improve pulse SNR in darker skin.
- **Offline operation:** deliver robust on-device inference with quality gating and asynchronous sync for low-bandwidth clinics [64].
- **Human factors:** embed pose/lighting guidance, motion tolerance, and transparent failure states to reduce unusable captures.

### Priorities for evaluation

- Spectrum-balanced, multi-site African validations powered for subgroup sensitivity/specificity in V/VI, with STARD-consistent methods, objective lighting (lux) and melanin measures alongside Fitzpatrick.
- Reference standards beyond clinic cuffs where feasible (e.g., ABPM/home BP) and pre-specified pairing windows to limit physiological drift [28].
- Head-to-head, multi-device experiments comparing on-device vs. cloud inference and quantifying bandwidth thresholds for feasibility/latency.
- Post-market fairness monitoring if deployed for any purpose.

### Bottom line

rPPG remains a promising route to scalable, contactless vital-sign capture. However, given the very low sensitivity in Fitzpatrick V–VI and bandwidth-sensitive feasibility observed here, algorithmic re-engineering, equity-centred validation, and robust offline operation are prerequisites before any screening use in similar settings [22, 18, 65].

## Data Availability

De-identified site-day internet speed aggregates underlying the figures and tables are provided in the Supplementary Materials. Raw rPPG signals were deleted upon completion of the study. Additional analysis code and aggregated files are available from the corresponding author upon reasonable request.

## Data availability

The datasets generated and analysed during the current study contain sensitive participant information and are not publicly available in accordance with participant consent agreements. De-identified summary data supporting the findings of this study are available from the corresponding author upon reasonable request. The underlying rPPG data streams were deleted after analysis and are therefore not available.

## Author contributions

**D.D.** conceived and designed the study, obtained ethics approvals (with support from D.P.), coordinated logistics, led participant recruitment and data collection in Nigeria, performed data cleaning and statistical analyses, prepared the figures, interpreted the results, and drafted the manuscript. **D.P.** supported pursuit of ethics approval, supervised the work, and provided critical revisions. Both authors approved the final manuscript and agree to be accountable for all aspects of the work.

## Acknowledgements

The authors thank the management and outpatient department staff of Sir Yahaya Memorial Hospital Birnin Kebbi, Martha Bamaiyi General Hospital Zuru, and Bamaiyi Sundu Memorial Medical Centre Senchi for their assistance in organizing participant recruitment and supporting data capture.

## References

[1] K. T. Mills, A. Stefanescu, J. He, The global epidemiology of hypertension, Nature Reviews Nephrology 16 (4) (2020) 223–237.

[2] World Health Organization, Hypertension, https://www.who.int/news-room/fact-sheets/detail/hypertension (2023).

[3] B. Zhou, P. Perel, G. A. Mensah, M. Ezzati, Global epidemiology, health burden and effective interventions for elevated blood pressure and hypertension, Nature Reviews Cardiology 18 (11) (2021) 785–802. doi: 10.1038/s41569-021-00559-8.

[4] S. Brouwers, I. Sudano, Y. Kokubo, E. M. Sulaica, Arterial hypertension, The Lancet 398 (10296) (2021) 249–261.

[5] National Institute for Health and Care Excellence, Hypertension in adults: diagnosis and management (nice guideline ng136), https://www.ncbi.nlm.nih.gov/books/NBK547161/ (2019).

[6] World Health Organization, First who report details devastating impact of hypertension and ways to stop it, https://www.who.int/news/item/19-09-2023-first-who-report-details-devastating-impact-of-hypertension-and-ways-to-story (2023).

[7] D. Adeloye, E. O. Owolabi, D. B. Ojji, A. Auta, M. T. Dewan, T. O. Olanrewaju, O. S. Ogah, C. Omoyele, N. Ezeigwe, R. G. Mpazanje, Prevalence, awareness, treatment, and control of hypertension in nigeria in 1995 and 2020: A systematic analysis of current evidence, The Journal of Clinical Hypertension 23 (5) (2021) 963–977.

[8] P. M. Kolo, Y. B. Jibrin, E. O. Sanya, M. Alkali, I. B. Peter Kio, R. K. Moronkola, Hypertension-related admissions and outcome in a tertiary hospital in northeast nigeria, International Journal of Hypertension 2012 (1) (2012) 960546.

[9] J. N. Kadima, B. Bavhure, J. D. Sepa, D. Murhura, Hypertensive urgencies or emergencies and co-morbidities in bukavu referral hospitals: clinical profile, management regimens, outcomes and drug related problems, Journal of Basic and Clinical Pharmacy 9 (1) (2018) 1.

[10] B.-M. Schmidt, S. Durao, I. Toews, C. M. Bavuma, A. Hohlfeld, E. Nury, J. J. Meerpohl, T. Kredo, Screening strategies for hypertension, Cochrane Database of Systematic Reviews (5) (2020).

[11] J. M. Guirguis-Blake, C. V. Evans, E. M. Webber, E. L. Coppola, L. A. Perdue, M. S. Weyrich, Screening for hypertension in adults: updated evidence report and systematic review for the us preventive services task force, JAMA 325 (16) (2021) 1657–1669.

[12] M. Boniol, T. Kunjumen, T. S. Nair, A. Siyam, J. Campbell, K. Diallo, The global health workforce stock and distribution in 2020 and 2030: a threat to equity and ‘universal’health coverage?, BMJ Global Health 7 (6) (2022) e009316.

[13] O. John, N. R. C. Campbell, T. M. Brady, M. Farrell, C. Varghese, A. Velazquez Berumen, L. A. Velez Ruiz Gaitan, N. Toffelmire, M. Ameel, M. Mideksa, The 2020 “who technical specifications for automated noninvasive blood pressure measuring devices with cuff”, Hypertension 77 (3) (2021) 806–812.

[14] W. Wang, A. C. d. Brinker, S. Stuijk, G. de Haan, Algorithmic principles of remote ppg, IEEE Transactions on Biomedical Engineering 64 (7) (2017) 1479–1491.

[15] P. V. Rouast, M. T. P. Adam, R. Chiong, D. Cornforth, E. Lux, Remote heart rate measurement using low-cost rgb face video: a technical literature review, Frontiers of Computer Science 12 (5) (2018) 858–872. doi:10.1007/s11704-016-6243-6.

[16] Y. Ba, Z. Wang, K. D. Karinca, O. D. Bozkurt, A. Kadambi, Overcoming difficulty in obtaining dark-skinned subjects for remote-ppg by synthetic augmentation, arXiv preprint arXiv:2106.06007 (2021).

[17] B. Kim, H. Kim, K. Kim, S. Kim, J. Kim, Learning not to learn: Training deep neural networks with biased data, in: Proceedings of the IEEE/CVF Conference on Computer Vision and Pattern Recognition, 2019, pp. 9012–9020.

[18] E. M. Nowara, D. McDuff, A. Veeraraghavan, A meta-analysis of the impact of skin tone and gender on non-contact photoplethysmography measurements, in: Proceedings of the IEEE/CVF Conference on Computer Vision and Pattern Recognition Workshops, 2020, pp. 284–285.

[19] Z. Zhang, J. M. Girard, Y. Wu, X. Zhang, P. Liu, U. Ciftci, S. Canavan, M. Reale, A. Horowitz, H. Yang, Multimodal spontaneous emotion corpus for human behavior analysis, in: Proceedings of the IEEE Conference on Computer Vision and Pattern Recognition, 2016, pp. 3438–3446.

[20] J. R. Estepp, E. B. Blackford, C. M. Meier, Recovering pulse rate during motion artifact with a multi-imager array for non-contact imaging photoplethysmography, in: 2014 IEEE International Conference on Systems, Man, and Cybernetics (SMC), 2014, pp. 1462–1469. doi:10.1109/SMC.2014.6974121.

[21] X. Liu, G. Narayanswamy, A. Paruchuri, X. Zhang, J. Tang, Y. Zhang, Y. Wang, S. Sengupta, S. Patel, D. McDuff, rppg-toolbox: Deep remote ppg toolbox, arXiv preprint (2022). arXiv:2210.00716. URL https://arxiv.org/abs/2210.00716

[22] E. Heiden, T. Jones, A. Brogaard Maczka, M. Kapoor, M. Chauhan, L. Wiffen, H. Barham, J. Holland, M. Saxena, S. Wegerif, Measurement of vital signs using lifelight remote photoplethysmography: results of the vision-d and vision-v observational studies, JMIR Formative Research 6 (11) (2022) e36340.

[23] NICE, Lifelight first for monitoring vital signs, London (2020).

[24] T. S. Ezeudu, T. J. Fadeyi, Examining the influence of infrastructure deficit on economic activities, education, and healthcare in rural areas of nigeria, Nnamdi Azikiwe Journal of Political Science 9 (1) (2024) 155–176.

[25] I. S. Abdulraheem, A. R. Olapipo, M. O. Amodu, Primary health care services in nigeria: Critical issues and strategies for enhancing the use by the rural communities, Journal of Public Health and Epidemiology 4 (1) (2012) 5–13.

[26] XIM Ltd., Lifelight – instructions for use (single product), https://xim-lifelight.github.io/ifu/ll-ifu/ll-singleproduct-ifu-c1.html, iFU Ref: REC-QMS-005, Revision 5; Date of issue: 06 Nov 2023 (2023). URL https://xim-lifelight.github.io/ifu/ll-ifu/ll-singleproduct-ifu-c1.html

[27] GSMA, The state of mobile internet connectivity 2023, London (2023).

[28] Ansi/aami/iso 81060-2:2019 non-invasive sphygmomanometers – part 2: Clinical investigation of intermittent automated measurement type (2019).

[29] P. M. Bossuyt, J. B. Reitsma, D. E. Bruns, C. A. Gatsonis, P. P. Glasziou, L. M. Irwig, J. G. Lijmer, D. Moher, D. Rennie, H. C. W. de Vet, Stard 2015: An updated list of essential items for reporting diagnostic accuracy studies, Radiology 277 (3) (2015) 826–832. doi:10.1148/radiol.2015151516. URL https://doi.org/10.1148/radiol.2015151516

[30] J. Tang, X. Li, J. Liu, X. Zhang, Z. Wang, Y. Wang, Camera-based remote physiology sensing for hundreds of subjects across skin tones, arXiv preprint arXiv:2404.05003 (2024).

[31] G. Boccignone, D. Conte, V. Cuculo, A. D’Amelio, G. Grossi, R. Lanzarotti, An open framework for remote-ppg methods and their assessment, IEEE Access 8 (2020) 216083–216103.

[32] L. D. van Putten, K. E. Bamford, Improving systolic blood pressure prediction from remote photoplethysmography using a stacked ensemble regressor, in: Proceedings of the IEEE/CVF Conference on Computer Vision and Pattern Recognition, 2023, pp. 5956–5963.

[33] C. El-Hajj, P. A. Kyriacou, A review of machine learning techniques in photoplethysmography for the non-invasive cuff-less measurement of blood pressure, Biomedical Signal Processing and Control (2020).

[34] N. Norori, Q. Hu, F. M. Aellen, F. D. Faraci, A. Tzovara, Addressing bias in big data and ai for health care: A call for open science, Patterns 2 (10) (2021).

[35] U. Pawar, D. O’Shea, S. Rea, R. O’Reilly, Explainable ai in healthcare, in: 2020 International Conference on Cyber Situational Awareness, Data Analytics and Assessment (CyberSA), 2020, pp. 1–2.

[36] P. Pirzada, A. Wilde, G. Doherty, D. Harris-Birtill, Remote photoplethysmography (rppg): A state-of-the-art review, medRxiv (2023) 2010–2023.

[37] F. Niel, M. Sahasrabudhe, V. Vives, L. Abensur Vuillaume, C. Gil-Jardine, C. Gerlier, Prospective clinical validation of a noncontact vital signs measurement smartphone application in emergency department, European Heart Journal 44 (Supplement 2) (2023) ehad655.2986. doi: 10.1093/eurheartj/ehad655.2986.

[38] T. A. Adisa, C. Mordi, A. R. Timming, Employment discrimination against indigenous people with tribal marks in nigeria: The painful face of stigma, Work, Employment and Society 38 (3) (2024) 787–808.

[39] A. Adeyemi, Facial scarification on ifé brass heads: An alternative hypothesis and its implications, Journal of Anthropology and Archaeology 3 (2) (2015) 21–36.

[40] E. O’Brien, N. Atkins, G. Stergiou, et al., European society of hypertension international protocol revision 2010 for the validation of blood pressure measuring devices in adults, Blood Pressure Monitoring 15 (1) (2010) 23–38. doi:10.1097/MBP.0b013e3283360e5c.

[41] Apple, ipad (8th generation) – technical specifications, https://www.apple.com/ipad-8th-generation/specs/, accessed: 2024-07-11 (2020).

[42] C. Sala, E. Santin, M. Rescaldani, F. Magrini, How long shall the patient rest before clinic blood pressure measurement?, American Journal of Hypertension 19 (7) (2006) 713–717.

[43] Medicines and Healthcare products Regulatory Agency, Public access registration database: Xim limited, mHRA Reference Number: 9515, Date Registered: 22 Jan 2020 (2020). URL https://pard.mhra.gov.uk/manufacturer-search/xim%20limited

[44] Companies House, Xim limited, https://find-and-update.company-information.service.gov.uk/company/03699022 (2024).

[45] L. D. van Putten, K. E. Bamford, I. Veleslavov, S. Wegerif, From video to vital signs: using personal device cameras to measure pulse rate and predict blood pressure using explainable ai, Discover Applied Sciences 6 (4) (2024) 184. doi:10.1007/s42452-024-05848-8.

[46] S. Sachdeva, Fitzpatrick skin typing: Applications in dermatology, Indian Journal of Dermatology, Venereology and Leprology 75 (2009) 93.

[47] W. Coleman, K. Mariwalla, P. Grimes, Updating the fitzpatrick classification: The skin color and ethnicity scale, Dermatologic Surgery 49 (8) (2023) 725–731.

[48] Federal Government of Nigeria, Nigeria data protection act, Federal Republic of Nigeria Official Gazette (2023).

[49] S. Bommareddy, J. A. Khan, R. Anand, A review on healthcare data privacy and security, in: Networking Technologies in Smart Healthcare, 2022, pp. 165–187.

[50] D. G. Altman, J. M. Bland, Measurement in medicine: The analysis of method comparison studies, Journal of the Royal Statistical Society. Series D (The Statistician) 32 (3) (1983) 307–317. doi:10.2307/2987937.

[51] V. Braun, V. Clarke, Using thematic analysis in psychology, Qualitative Research in Psychology 3 (2) (2006) 77–101.

[52] National Health Research Ethics Committee of Nigeria, National code of health ethics, Federal Ministry of Health, Department of Health Planning and Research (2007).

[53] M. D. E. Goodyear, K. Krleza-Jeric, T. Lemmens, The declaration of helsinki, BMJ (2007).

[54] C. J. Clopper, E. S. Pearson, The use of confidence or fiducial limits illustrated in the case of the binomial, Biometrika 26 (4) (1934) 404–413. URL http://www.jstor.org/stable/2331986

[55] D. S. Wilson, Confidence intervals for motion and deformation of the juan de fuca plate, Journal of Geophysical Research: Solid Earth 98 (B9) (1993) 16053–16071. arXiv:https://agupubs.onlinelibrary.wiley.com/doi/pdf/10.1029/93JB01227, 10.1029/93JB01227. URL https://agupubs.onlinelibrary.wiley.com/doi/abs/10.1029/93JB01227

[56] T. pandas development team, pandas-dev/pandas: Pandas, Zenodo (2020). doi:10.5281/zenodo.3509134. URL https://doi.org/10.5281/zenodo.3509134

[57] C. R. Harris, K. J. Millman, S. J. van der Walt, R. Gommers, P. Virtanen, D. Cournapeau, E. Wieser, J. Taylor, S. Berg, N. J. Smith, R. Kern, M. Picus, S. Hoyer, M. H. van Kerkwijk, M. Brett, A. Haldane, J. F. del Ŕıo, M. Wiebe, P. Peterson, P. Gérard-Marchant, K. Sheppard, T. Reddy, W. Weckesser, H. Abbasi, C. Gohlke, T. E. Oliphant, Array programming with NumPy, Nature 585 (2020) 357–362. doi: 10.1038/s41586-020-2649-2.

[58] P. Virtanen, R. Gommers, T. E. Oliphant, M. Haberland, T. Reddy, D. Cournapeau, E. Burovski, P. Peterson, W. Weckesser, J. Bright, S. J. van der Walt, M. Brett, J. Wilson, K. J. Millman, N. Mayorov, A. R. J. Nelson, E. Jones, R. Kern, E. Larson, C. Carey, İ. Polat, Y. Feng, E. W. Moore, J. VanderPlas, D. Laxalde, J. Perktold, R. Cimrman, I. Henriksen, E. A. Quintero, C. R. Harris, A. M. Archibald, A. H. Ribeiro, F. Pedregosa, P. van Mulbregt, SciPy 1.0 Contributors, SciPy 1.0: fundamental algorithms for scientific computing in python, Nature Methods 17 (2020) 261–272. doi:10.1038/s41592-019-0686-2.

[59] S. Seabold, J. Perktold, statsmodels: Econometric and statistical modeling with python (2010) 57–61. URL https://conference.scipy.org/proceedings/scipy2010/seabold.html

[60] J. D. Hunter, Matplotlib: A 2d graphics environment, Computing in Science & Engineering 9 (3) (2007) 90–95. doi:10.1109/MCSE.2007.55.

[61] M. L. Waskom, seaborn: statistical data visualization (2021). doi:10.21105/joss.03021.

[62] T. Rietveld, R. van Hout, The paired t test and beyond: Recommendations for testing the central tendencies of two paired samples in research on speech, language and hearing pathology, Journal of Communication Disorders 69 (2017) 44–57.

[63] S. E. Maxwell, Sample size and multiple regression analysis, Psychological Methods 5 (4) (2000) 434.

[64] P. P. Sust, O. Solans, J. C. Fajardo, M. M. Peralta, P. Rodenas, J. Gabaldà, L. G. Eroles, A. Comella, C. V. Muñoz, J. S. Ribes, Turning the crisis into an opportunity: digital health strategies deployed during the covid-19 outbreak, JMIR Public Health and Surveillance 6 (2) (2020) e19106.

[65] L. Lu, J. Zhang, Y. Xie, F. Gao, S. Xu, X. Wu, Z. Ye, Wearable health devices in health care: narrative systematic review, JMIR mHealth and uHealth 8 (11) (2020) e18907.

